# Atrial and Biventricular Dysfunction Persists after Catheter Ablation and is a Factor of Risk for Post-procedure Atrial Fibrillation Recurrence in Patients with Preserved Left Ventricular Ejection Fraction

**DOI:** 10.1101/2025.09.21.25336292

**Authors:** Radu Tanacli, Emanuel Heil, Matthias Bock, Nikolaos Dagres, Babken Asatryan, Fabian Bartels, Volkmar Falk, Gerhard Hindricks, Jin-Hong Gerds-Li, Sebastian Kelle, Felix Hohendanner

## Abstract

**Background:** Catheter ablation (CA) is the leading rhythm control strategy for atrial fibrillation (AF). Despite its effectiveness, AF recurrences, driven by underlying structural and electrical remodeling in both the atria and the ventricle, remain prevalent. Cardiac magnetic resonance (CMR) imaging has emerged as a key modality for assessing myocardial remodeling and potentially identifying predictors of AF recurrence. This study aimed to investigate atrial and biventricular remodeling following CA of AF and its impact on AF recurrence.

**Methods and Results:** Fifty patients with AF (52% male, age 68±10 years) and twenty healthy controls (45% male, age 68±10 years) underwent CMR at least three months following CA. Recurrence of AF was evaluated over 1-year follow-up and defined as at least two unsuccessful attempts to restore SR. Atrial function was measured using phasic volumetric and strain parameters. Ventricular function was evaluated through volumetry, global left ventricular (LV) and right ventricular (RV) strain and LV structure through parametric mapping (native T1/T2 mapping and extracellular volume (ECV)). Patients with AF recurrence at 1 year showed significantly impaired LA active emptying function (14±12% vs 26±14%, P=0.009) and strain (−4.5±4.3% vs −10.6±6.8%, P=0.004), prolongation of both LV T1 (1040±55ms vs 985±43ms, P=0.007) and T2 (53.6±1.6ms vs 50.6±2.5ms, P=0.002) times and elevated LV ECV (28.0±3.0% vs 26.2±1.7%, P=0.027) compared with those without recurrence. A combined model incorporating LA active strain, LV T1 native, ECV and T2 further improved prediction of AF recurrence at 1-year follow-up (HR: 17.70, 95% CI=3.89 - 80.61, log-rank P value <0.001 for cut-off values of LA Active Strain > −8%, T1 > 1017.3ms, ECV > 27% and T2 > 52.4ms). RV free wall longitudinal strain was more negative in AF non-recurrence group compared with controls but not in the AF recurrence group, suggesting the compensatory importance of RV function in post-procedural hemodynamic recovery.

**Conclusion:** AF recurrence after CA appears linked to persistent atrial dysfunction and ventricular remodelling, potentially fostering a pro-arrhythmic milieu. Incorporating CMR-derived measures into AF care might help improve personalized risk stratification for CA outcomes.

## Background

Catheter ablation (CA) has emerged as the cornerstone rhythm control strategy for atrial fibrillation (AF), in patients with preserved and reduced left ventricular function alike. ^1^ By electrically isolating arrhythmogenic foci within the pulmonary veins, CA offers superior rhythm control compared to antiarrhythmic drugs ^2^, the use of which is limited by their suboptimal efficacy and broad spectrum of side effects. Especially in patients with structurally normal hearts, CA not only restores sinus rhythm but also interrupts the vicious cycle of atrial and ventricular remodelling and a missing left atrial kick, mitigating the risk of heart failure. ^3^ Despite advancements in electro-anatomical mapping and implementation of novel ablation methods, like the introduction of pulsed-field ablation, a significant proportion of patients still experience AF recurrence, necessitating repeat interventions. These cases underscore the complex and multifaceted pathophysiology of AF, which extends beyond electrical disarray to involve profound structural and molecular remodelling. AF reflects a dynamic interplay between atrial and ventricular pathology, driven by mechanisms such as ion channel dysfunction, aberrant calcium handling, oxidative stress, inflammation, and interstitial fibrosis. ^4,5^ Furthermore, the presence of subclinical structural cardiac abnormalities—such as left atrial enlargement or diffuse ventricular fibrosis—often precedes overt AF and signifies an advanced myocardial remodelling process. ^6^

At the cellular level, mitochondrial dysfunction, extracellular matrix dysregulation, and chronic inflammatory signalling converge to create a substrate conducive to AF initiation and maintenance. Systemic comorbidities, including hypertension, obesity, and metabolic syndrome, exacerbate these pathophysiological changes, perpetuating a cycle of myocardial dysfunction and arrhythmogenesis. The ability of CA to reverse or halt these structural changes, particularly atrial fibrosis and inflammation, remains a pivotal determinant of long-term success.^7,8^ Conversely, the persistence of these structural abnormalities post-ablation can sustain a pro-arrhythmic substrate, increasing the likelihood of AF recurrence. This highlights the critical need for targeted strategies to address remodelling comprehensively as part of AF management.

The advent of cardiac magnetic resonance (CMR) imaging has transformed the evaluation of myocardial remodelling, offering unparalleled insights into both atrial and ventricular tissue architecture and function. Beyond its established role in quantifying the volume and function of all four cardiac chambers ^9^ and replacement fibrosis using late gadolinium enhancement (LGE) ^10^ with high accuracy, CMR enables advanced parametric mapping to assess myocardial tissue properties such as fibrosis, oedema, and fat infiltration. ^11–13^ This technology also facilitates detailed imaging of the right atrium and right ventricle, structures often overlooked in standard echocardiographic assessments despite their integral roles in AF pathophysiology. ^14^

In this study, we aimed to leverage CMR to provide a comprehensive assessment of cardiac remodelling across all four chambers following CA. By characterizing the long-term structural and functional consequences of the disease and its treatment, we seek to identify predictors of AF recurrence and further elucidate the pathophysiological mechanisms underlying AF recurrence.

## Methods

### Study population

A total of 50 patients who underwent first-time radiofrequency catheter ablation (CA) for paroxysmal atrial fibrillation between January 2022 and October 2023 were retrospectively included. The study received ethics approval from the local Institutional Review Board (Charité – Universitätsmedizin Berlin; EA1/324/21). All research complied with the Belmont Report principles and the Declaration of Helsinki. Because all procedures and cardiac magnetic resonance (CMR) examinations were clinically indicated, no study-specific interventions were performed. Written informed consent for participation and for the use of clinical data for research (including retrospective access to clinical records) was obtained from all included patients at the time of clinical screening. Clinical records and imaging metadata were accessed for research purposes between June and November 2023; during chart abstraction, authorised study personnel accessed the electronic health record, which contained direct identifiers. The analysis dataset used for this study was de-identified prior to analysis, and no author had access to direct identifiers after dataset creation. All data were de-identified prior to analysis; no directly identifying information is presented. As a matched control group, 20 individuals without AF who underwent clinical CMR but no CA from a local database were included; some of these control subjects were included in previous studies.^15–18^ Control-group records were accessed in the same period under the same approval. All patients underwent standard pre-procedural evaluation, including transoesophageal echocardiography to exclude left atrial thrombus and computed tomography or magnetic resonance imaging to assess left atrial anatomy. Antiarrhythmic drugs were discontinued for at least five half-lives (amiodarone six weeks) per institutional protocol. Procedures were performed under conscious sedation or general anaesthesia with periprocedural anticoagulation per institutional protocol. This observational study did not prospectively assign participants to interventions and therefore does not meet the WHO/ICMJE definition of a clinical trial; trial registration was not applicable. Reporting follows STROBE recommendations.

### Catheter Ablation Procedure. Electro-anatomical Mapping and LA Pressure Measurement

The maintenance of SR has been assessed by monthly 24hrs Holter evaluation performed in the OP clinic and by telephone interviews regarding symptoms. A three-dimensional electroanatomical mapping system (Carto 3; Biosense Webster, Diamond Bar, CA) was utilized for all procedures. Upon standardized vascular access under conscious sedation using propofol, a decapolar catheter was positioned in the coronary sinus. A high-density mapping catheter (e.g., Pentaray or Octaray) was advanced into the left atrium upon transseptal puncture using a Brockenbrough needle approach under fluoroscopic and, on a case-by-case basis, echocardiographic guidance. Heparin was administered to maintain an activated clotting time of 300–350 seconds throughout the procedure. High-resolution voltage and activation maps of the left atrium were created during sinus rhythm (i.e., if patients were in AF at the procedure onset, electrical cardioversion was performed). Bipolar electrograms were recorded with a filter setting of 30–500 Hz, and a voltage range of 0.05–0.5 mV was used to define low-voltage areas. The electroanatomical maps were acquired with a density of at least 1,000 points to ensure high map accuracy.

### Follow-up and Assessment of Procedural Success

Procedural success was characterized as the absence of any atrial arrhythmia following a 3-month blanking period. After the procedure, patients were monitored in outpatient clinic visits at 1, 3, 6, and 12 months. Patients underwent 24-hours Holter monitoring at 3, 6, and 12-month intervals. At the time of discharge and each clinical visit, those experiencing recurrent symptoms were advised to contact the arrhythmia unit for prompt evaluation and use of an event monitor. Recurrence was identified as any documented sustained atrial arrhythmia lasting more than 30 seconds, in alignment with established consensus guidelines ^19,20^ or any AF recurrence documentation by a physician. In this study, we considered AF recurrence patients as those patients who experienced episodes of AF after undergoing two CA procedures.

### CMR Protocol. Image acquisition

A post-procedure CMR was performed at least 3 months after the procedure. All exams were performed on a clinical 1.5T MRI system (Ambition, Philips Healthcare, Best, The Netherlands) using a specific cardiac 32-element phased array coil. Cine short-axis, vertical long-axis and horizontal long-axis steady-state free precession (SSFP) sequences using a retrospectively gated cine-CMR. T1 Native and at 15 min post-contrast maps were acquired using a modified Look-Locker (MOLLI) 5s(3s)3s-scheme. ^21^ Imaging parameters were as follows: Acquired voxel size = 2.0 × 2.0 × 10mm^3^, reconstructed voxel size = 0.5 × 0.5 × 10mm^3^, balanced SSFP readout, flip angle = 35°, parallel imaging (SENSE) factor = 2 and effective inversion times between 150 and 3382ms. T2-mapping was performed before administration of contrast media using a black-blood-prepared, navigator-gated, free-breathing hybrid gradient (echo planar imaging, EPI) and a spin-echo multi-echo sequence (GraSE), as described previously, with the following typical imaging parameters: TR = 1 heartbeat, 9 echoes (TE1 = 15ms, delta TE = 7.7ms), FA 90°, parallel imaging (SENSE = 2), EPI factor = 7, black-blood prepulse and breath-hold (duration of the sequence about 14 s).

### CMR Protocol. Image analysis

All image analyses were performed using Medis Suite version 3.2 (Medis Medical Imaging Systems bv Leiden, The Netherlands). Biventricular volumetry was evaluated using the most recent recommendations of the Society for Cardiovascular Magnetic Resonance, SCMR. LV and global longitudinal and circumferential strain and RV global longitudinal and free wall strain were measured following the algorithms we described previously. ^18,22^

LA maximal and minimal volumes were first established visually in long-axis SSFP cine 4Ch and 2Ch views, respectively and manually assigned. All values were averaged between those obtained in 4Ch and 2Ch images, respectively. Atrial volumetric and strain curves were recomposed and all atrial phasic parameters were derived, as previously described^23^: LA Max, maximal LA volume; LA Min, minimal LA volume; LApreA, volume at atrial diastasis) (**Figure 1**), LA emptying fraction (LAMax−LAMin)/LAmax, passive LA emptying fraction (LAMax− LApreA)/LAMax, active LA emptying fraction (LApreA−LAmin)/LAMax, as previously described.^23^ Subsequently, these contours were further exported to a FT platform, QStrain RE version 2.0 and automatically propagated over the entire cardiac cycle. In a similar fashion, LA phasic strain was derived using LA Strain Max, Min and LApreA strain at atrial diastasis. ^24^ Similarly, the values were averaged between those obtained in 4Ch and 2Ch images, respectively.

**Figure 1.**
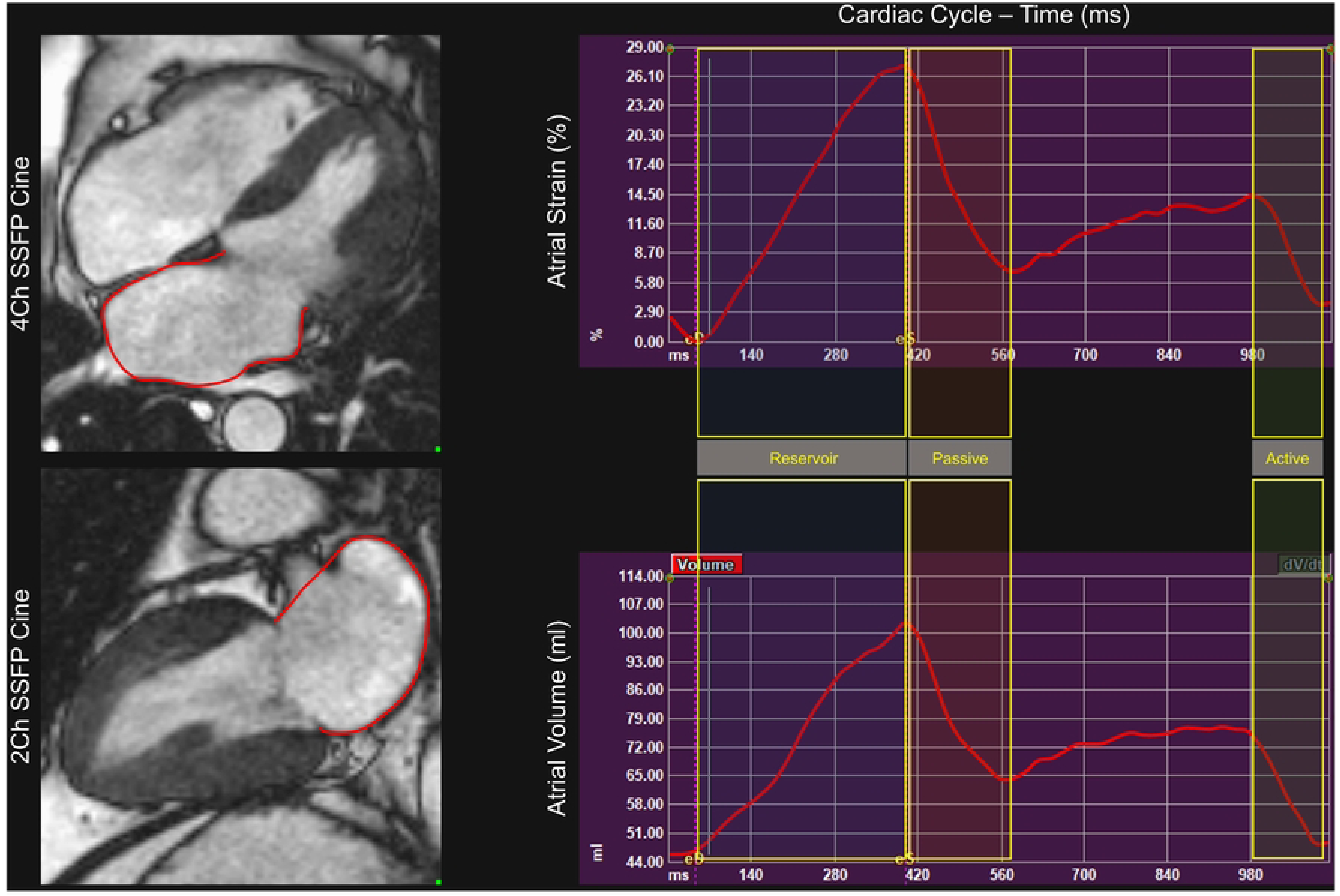
Phasic left atrial (LA) strain and volume. Contours are drawn on both 4-chamber (4Ch) and 2-chamber (2Ch) steady-state free precession (SSFP) cine images at the points of maximal and minimal volume and propagated throughout the cardiac cycle. Three phases of atrial physiology are depicted: (1) Reservoir phase, representing LA filling between minimal volume (corresponding to mitral valve closure at the end of left ventricular diastole) and maximal volume; (2) Passive emptying phase, representing LA emptying from mitral valve opening to atrial diastasis; and (3) Active emptying phase, representing LA emptying during atrial contraction, from the onset of the P wave on the ECG to mitral valve closure. LA phasic volumes and strains are calculated for each of these three phases.

### Echocardiography

A routine Doppler echocardiography exam was performed on all patients and controls included in this study, contemporary with the CMR scan. For our analysis, we focused on the parameters that characterized the LV diastolic function: E – early, A – atrial (late) diastolic transmitral flow velocities and E/e’ ratio, where e’ represents that early diastolic tissue displacement velocity of para-mitral LV segments, as an imaging approximation of LV filling pressure. ^25^ All echocardiography exams were performed on an EPIQ 7 ultrasound system (Philips Healthcare, Hamburg, Germany).

### Statistics

Data were analysed using dedicated statistical packages in Python (Python Software Foundation, version 3.11), R (R Foundation for Statistical Computing, version 4.2.2), and GraphPad Prism (GraphPad Software, version 9, San Diego, CA). pROC package was used in R for ROC curve analysis and DeLong’s test, and scikit-learn module was used in Python for AUC calculation and logistic regression. The Seaborn package (version 0.12.2) in Python was additionally used for graphs. All variables were tested for normal distribution using the Kolmogorov-Smirnov test. The equality of variances between subgroups was tested with Brown-Forsythe test. ANOVA one-way or non-parametric equivalent test were used to compare continuous variables between three or more groups and a Tukey post-hoc test with Bonferroni correction to compare further the mean between each two of these groups. ROC analysis followed by determination of specific area under curve (AUC) and Youden’s index calculation was used to determine the optimal cut-off value, sensitivity and specificity. Logistic regression was performed to assess the relationship between the variables T1 Native, LV ECV, and T2, with the outcome variable being the true label. The logistic regression model was then extended by incorporating the LA Active Strain as an additional predictor, creating a combined model to improve the prediction of the outcome. To compare the AUCs of the two individual parameters with the combined model and assess the statistical significance of ROC analyses, DeLong’s test was used. Results are presented as mean ± SD. Values of p<0.05 were considered statistically significant.

## Results

### Study population

Table 1 presents complete demographic, comorbidities, and baseline blood test results. Of the 50 patients with AF included in the study, 14 were considered to have AF recurrence, and 36 were considered arrhythmia-free at one-year follow-up.

**Table 1.** Population Characteristics in 3 subgroups: AF patients with recurrence, AF patients with no recurrence and Control. Demographics, Comorbidities. Blood Test Results.

|  |  |  |  |  |  |  |  |
| --- | --- | --- | --- | --- | --- | --- | --- |
|  | AF-recurrence<br>N=14 | AF-no recurrence<br>N=36 | Control<br>N=20 | P<br>AF rec vs no rec | P<br>AF rec vs Control | P<br>AF no rec vs Control | P<br>ANOVA/Chi sq |
| **Clinical Characteristics** |  |  |  |  |  |  |  |
| Age, y | 67±12 | 69±10 | 69±10 | 0.91 | 0.88 | 0.99 | 0.88 |
| Sex, males | 8(57) | 18(50) | 11(45) | 0.76 | 0.99 | 0.78 | 0.26 |
| BMI | 24±3 | 29±6 | 25±3 | **0.007** | 0.35 | **0.007** | **0.015** |
| **Comorbidities** |  |  |  |  |  |  |  |
| Hypertension | 9 (64) | 22(61) | 9(45) | 0.99 | 0.25 | 0.32 | 0.42 |
| CAD Obstructive | 0(0) | 0(0) | 0(0) | 0.99 | 0.99 | 0.99 | 0.99 |
| CAD Non-obstructive | 1(7) | 4(11) | 1(5) | 0.99 | 0.99 | 0.64 | 0.72 |
| Previous MI | 0(0) | 0(0) | 0(0) | 0.99 | 0.99 | 0.99 | 0.99 |
| Diabetes | 2(14) | 4(11) | 1(5) | 0.99 | 0.56 | 0.64 | 0.64 |
| Dyslipidemia | 3(21) | 7(19) | 5(25) | 0.99 | 0.99 | 0.74 | 0.89 |
| Asthma/COPD | 1(7) | 4(11) | 2(10) | 0.99 | 0.99 | 0.99 | 0.92 |
| Sleep Apnoea | 3(21) | 9(25) | 4(20) | 0.99 | 0.99 | 0.75 | 0.90 |
| **Blood Tests** |  |  |  |  |  |  |  |
| Hb (g/dl) | 13.7±1.4 | 14.7±1.3 | 13.9±1.1 | 0.11 | 0.91 | 0.16 | 0.07 |
| Htc (%) | 41±4 | 43±3 | 42±5 | 0.08 | 0.54 | 0.35 | 0.32 |
| WBC (K/ul) | 6.5±1.8 | 6.6±1.6 | 6.1±1.6 | 0.98 | 0.77 | 0.56 | 0.57 |
| HbA1C (%) | 5.7±0.4 | 5.6±0.4 | 5.4±0.5 | 0.8 | 0.41 | 0.72 | 0.43 |
| Glucose (mg/dl) | 106±18 | 110±28 | 107±14 | 0.51 | 0.87 | 0.63 | 0.70 |
| Urea (mg/dl) | 37.8±6.6 | 39.9±10.3 | 31.0±4.8 | 0.75 | 0.07 | **0.002** | **0.003** |
| Creatinine (mg/dl) | 1.04±0.12 | 1.04±0.22 | 0.88±0.19 | 0.99 | 0.10 | **0.035** | **0.027** |
| eGFR (ml/min/1.72m2) | 68±12 | 75±18 | 80±10 | 0.33 | 0.06 | 0.56 | 0.08 |
| Triglycerides | 90±25 | 170±150 | 130±80 | 0.13 | 0.60 | 0.49 | 0.146 |
| Total Cholesterol | 170±22 | 167±47 | 203±33 | 0.97 | 0.06 | **0.012** | **0.010** |
| HDL Cholesterol | 59±20 | 48±9 | 66±25 | 0.28 | 0.59 | **0.012** | **0.016** |
| LDL Cholesterol | 99±28 | 97±34 | 133±39 | 0.99 | **0.041** | **0.008** | **0.006** |
| Albumin | 4.0±0.4 | 4.2±0.2 | 4.4±0.2 | 0.88 | 0.26 | 0.44 | 0.71 |
| TSH | 1.14±0.89 | 1.55±0.61 | 2.15±1.70 | 0.67 | 0.11 | 0.28 | 0.10 |
| log NTpro-BNP | 2.97±0.35 | 2.80±0.56 | 1.84±0.32 | 0.50 | **<0.001** | **<0.001** | **<0.001** |
| Abbreviations: BMI body mass index, CAD coronary artery disease, COPD chronic obstructive pulmonary disease, Hb hemoglobin, Htc hematocrit, HbA1C glycosylated hemoglobin, eGFR estimated glomerular filtration rate, TSH thyroid-stimulating hormone, NTpro-BNP N-Terminal pro-B-Type natriuretic peptide |  |  |  |  |  |  |  |

We compared the following three groups for the present analysis: AF patients with recurrence (AF-r), AF patients without recurrence (AF-nr) and Control. The groups were equivalent with regard to age and sex distribution and the AF-nr subgroup had comparatively a slightly higher BMI. Controls tended to have lower creatinine levels and a better lipid profile than the AF patients. As expected, the Control group had lower NT-proBNP compared with both AF groups. There was no difference in comorbidities or laboratory results between AF-r and AF-nr groups.

### Atrial remodeling post AF

We investigated LA remodeling post-CA procedure using several methods and complete numerical details are presented in **Table 2**. Of note, all LA phasic volumes, reservoir, passive and active emptying fractions and strain were impaired in both AF groups compared with Control. (**Figure 2 A, B, C**) LA volume assessed by echocardiography and LA min assessed CMR was larger in the AF-r than in the AF-nr group. Only the active EF and strain were decisively different between AF-r and AF-nr groups. (Active LA EF: 14±12% vs 26±14%, P=0.009, Active LA Strain: −4.5±4.3% vs −10.6±6.8%, P=0.004). Reservoir EF was also reduced in AF-r compared with AF-nr (31±15% vs 41±14%, P=0.034), but this likely represented more an effect of differences in active EF as the passive EF was not different. With regard to the electro-anatomical mapping performed, there was no difference in the extent of low-voltage areas or percentages. Similarly, the atrial pressures measured during the CA procedure were equivalent between these two groups. For RA, there was no difference in volumes or phasic strain (**Figure 2 D, E, F**).

**Table 2.** Left Atrium Remodeling in 3 subgroups: AF patients with recurrence. AF patients with no recurrence and Control. Catheter-measured LA pressures. Electro-anatomical Mapping. Echocardiography and Cardiac Magnetic Resonance Parameters.

|  |  |  |  |  |  |  |  |
| --- | --- | --- | --- | --- | --- | --- | --- |
|  | AF-recurrence<br>N=14 | AF-no recurrence<br>N=36 | Control<br>N=20 | P<br>AF rec vs non-rec | P<br>AF rec vs Control | P<br>AF non-rec vs Control | P<br>ANOVA |
| **LA pressure** |  |  |  |  |  |  |  |
| LAP (a-wave), mmHg | 9±2 | 12±6 |  | 0.42 |  |  |  |
| LAP (v-wave), mmHg | 11±2 | 13±8 |  | 0.64 |  |  |  |
| LAP (mean), mmHg | 8±2 | 9±4 |  | 0.54 |  |  |  |
| LAP/LAV, mmHg*m2/ml | 0.12±0.02 | 0.29±0.17 |  | **0.021** |  |  |  |
| **EA mapping** |  |  |  |  |  |  |  |
| total area (cm<sup>2</sup>; Carto) | 196±60 | 171±62 |  | 0.22 |  |  |  |
| posterior wall total (cm<sup>2</sup>; Carto) | 25±13 | 30±17 |  | 0.35 |  |  |  |
| roof total (cm<sup>2</sup>; Carto) | 16±6 | 18±10 |  | 0.48 |  |  |  |
| anterior wall total (cm<sup>2</sup>; Carto) | 29±13 | 32±18 |  | 0.67 |  |  |  |
| posterior wall low voltage (cm<sup>2</sup>; Carto) | 2.9±6.1 | 5.0±10.3 |  | 0.83 |  |  |  |
| roof low voltage (cm<sup>2</sup>; Carto) | 4.6±7.6 | 4.2±6.6 |  | 0.87 |  |  |  |
| anterior wall low voltage (cm<sup>2</sup>; Carto) | 6.7±10.3 | 6.6±11.7 |  | 0.86 |  |  |  |
| global low voltage (%) | 18.9±25.7 | 15.4±21.2 |  | 0.63 |  |  |  |
| **CMR** |  |  |  |  |  |  |  |
| LAV Max | 65±25 | 53±14 | 40±8 | 0.07 | **<0.001** | **0.009** | **<0.001** |
| LAV Min | 46±27 | 32±14 | 15±3 | **0.026** | **<0.001** | **0.001** | **<0.001** |
| Reservoir LA EF, % | 31±15 | 41±14 | 63±4 | **0.034** | **<0.001** | **<0.001** | **<0.001** |
| Reservoir LA Strain, % | 16.1±8.7 | 22.7±10.8 | 33.1±7.6 | 0.09 | **<0.001** | **0.001** | **<0.001** |
| Passive LA EF, % | 21.4±9.4 | 22.2±6.4 | 29.1±7.3 | 0.93 | **0.012** | **0.004** | **0.002** |
| Passive LA Strain, % | -9.4±3.4 | -11.2±4.4 | -16.7±5.8 | 0.48 | **<0.001** | **<0.001** | **<0.001** |
| Active LA EF, % | 14±12 | 26±14 | 48±5 | **0.009** | **<0.001** | **<0.001** | **<0.001** |
| Active LA Strain, % | -4.5±4.3 | -10.6±6.8 | -17.3±3.9 | **0.004** | **<0.001** | **<0.001** | **<0.001** |

Abbreviations: LAP left atrial pressure, LAV left atrial volume, EA electro-anatomical, CMR cardiac magnetic resonance, EF ejection fraction

**Figure 2.**
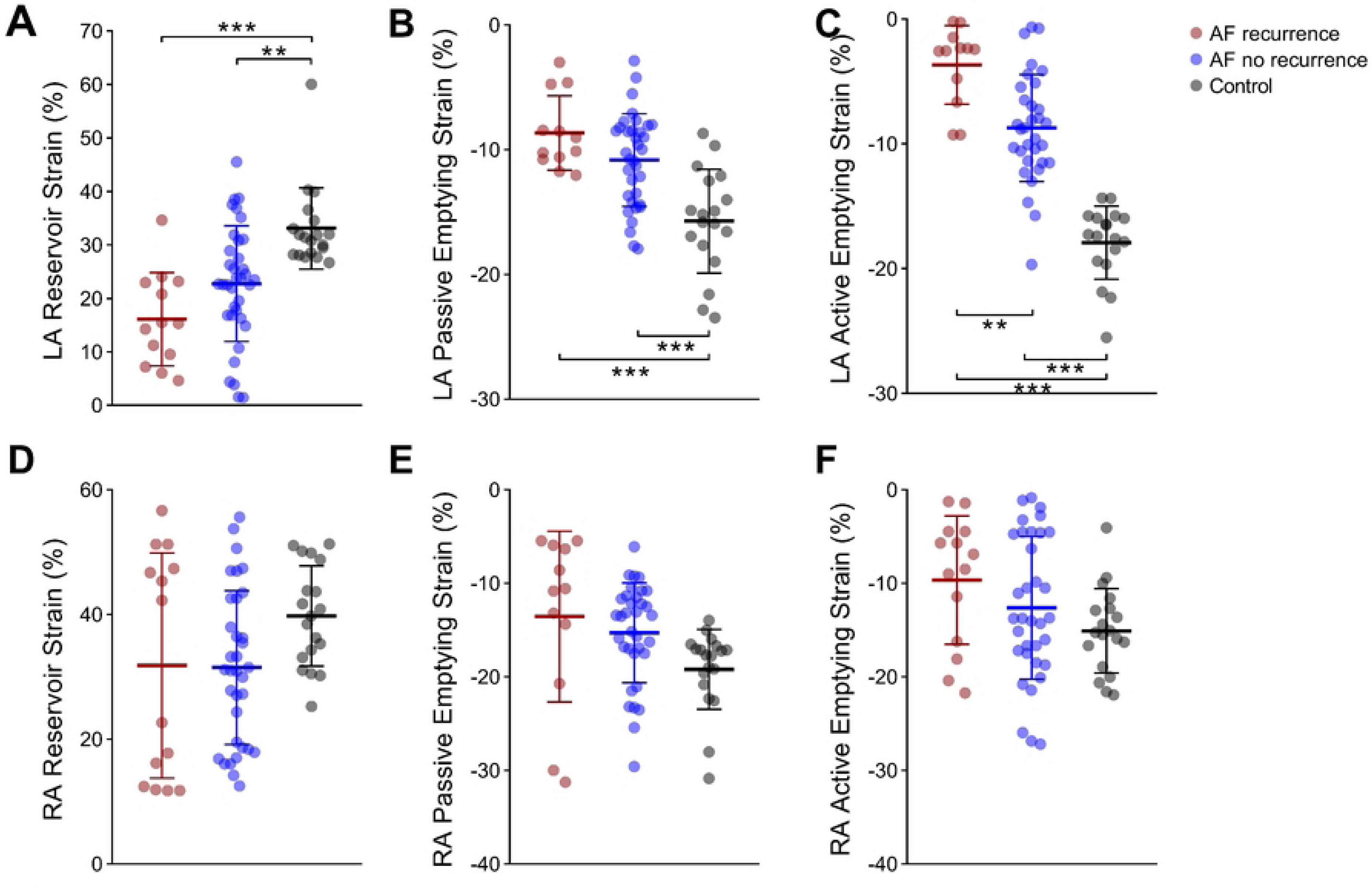
Phasic atrial strain analysis illustrating reservoir, passive, and active strains for the left atrium (Panels **A**, **B**, **C**) and the right atrium (Panels **D**, **E**, **F**) across three groups: AF patients with recurrence post-RFCA (red dots), AF patients without recurrence post-RFCA (blue dots), and controls (black dots). Left atrial phasic strains: reservoir strain (**A**), passive strain (**B**), and active strain (**C**), demonstrate persistent atrial dysfunction in AF compared with controls. LA Active Strain demonstrates differences between AF patients with recurrence vs AF patients with no recurrence. No similar differences are observed for the RA (Panels **D**, **E**, **F**). In all panels, phasic strains are presented as mean ± standard error, stratified by group. Significant differences between groups (p < 0.05) are indicated as follows: * p<0.05, ** p<0.01, *** p<0.001.

### Left ventricular remodeling

Next, we analyzed the differences in LV remodeling between the 3 groups with CMR Doppler echocardiography (details given in **Table 3** and illustrated in **Figure 3 A-F**). There was no difference in LV volumetric assessment between groups, but LV mass was increased in AF groups compared with Control (76±37g/m^2^ vs 71±31g/m^2^ vs 42±15g/m^2^, P_ANOVA_ = 0.001). However, this was not different between AF-r and AF-nr (P=0.87) (**Figure 3 D**). Functionally, LV EF was not different between groups, but there was an impairment in both AF groups compared with Control for longitudinal (−20.3±5.6% vs −20.5±3.7% vs −25.9±3.2%, P_ANOVA_ < 0.001) and circumferential (−30.3±8.7% vs −29.8±5.8% vs −34.7±5.8%, P_ANOVA_ = 0.029) global LV strain. (**Figure 3 A-C**). On parametric mapping imaging, T1 (1040±55ms vs 985±43ms vs 972±29ms, P_ANOVA_ = 0.001) and T2 (53.6±1.6ms vs 50.6±2.5ms vs 50.7±2.0ms, P_ANOVA_ = 0.006) times were longer in AF-r than AF-nr or Control, but not different between AF-nr and Control. ECV assessment requires contrast administration, and therefore, these were available only in patients with AF where values were higher in AF-r than in the AF-nr group (28.0±3.0% vs 26.2±1.7%, P=0.027).

**Table 3.** Left Ventricular Remodeling in 3 subgroups: AF patients with recurrence. AF patients with no recurrence and Control. Echocardiography and Cardiac Magnetic Resonance Parameters.

|  |  |  |  |  |  |  |  |
| --- | --- | --- | --- | --- | --- | --- | --- |
|  | AF-recurrence<br>N=14 | AF-no recurrence<br>N=36 | Control<br>N=20 | P<br>AF rec vs non-rec | P<br>AF rec vs Control | P<br>AF non-rec vs Control | P<br>ANOVA |
| **<u>Left Ventricle</u>** |  |  |  |  |  |  |  |
| **Echocardiography** |  |  |  |  |  |  |  |
| E/e' | 10.6±4.2 | 10.8±3.6 | 10.1±3.7 | 0.98 | 0.95 | 0.86 | 0.87 |
| e' (cm/s) | 8.2±1.9 | 8.6±2.9 | 8.4±2.2 | 0.86 | 0.99 | 0.95 | 0.89 |
| E Max (cm/s) | 83.7±24.3 | 85.4±22.0 | 64.7±16.9 | 0.97 | 0.10 | **0.025** | 0.029 |
| A Max (cm/s) | 36.5±17.3 | 53.9±15.1 | 71.4±18.1 | **0.031** | **<0.001** | **0.014** | <0.001 |
| Deceleration Time (s) | 0.19±0.07 | 0.22±0.07 | 0.19±0.05 | 0.49 | 0.97 | 0.62 | 0.31 |
| E/A | 3.00±2.01 | 1.74±0.88 | 0.93±0.20 | **0.017** | **<0.001** | 0.102 |  |
| IVSd, cm | 1.2±0.3 | 1.2±0.1 | 1.1±0.1 | 0.99 | 0.41 | 0.35 | 0.32 |
| LVIDd, cm | 4.8±0.8 | 5.0±0.5 | 4.6±0.4 | 0.70 | 0.62 | 0.15 | 0.17 |
| LVPWd, cm | 1.1±0.3 | 1.0±0.2 | 1.0±0.1 | 0.15 | 0.25 | 0.99 | 0.15 |
| **CMR** |  |  |  |  |  |  |  |
| EDV, ml/m2 | 73±10 | 73±14 | 78±13 | 0.99 | 0.55 | 0.38 | 0.38 |
| ESV, ml/m2 | 30±7 | 30±8 | 29±7 | 0.99 | 0.98 | 0.90 | 0.91 |
| SV, ml/m2 | 43±7 | 42±9 | 47±6 | 0.98 | 0.25 | 0.08 | 0.09 |
| CI, l/min/m2 | 2.85±0.70 | 2.79±0.64 | 2.82±0.50 | 0.96 | 0.99 | 0.99 | 0.96 |
| EF, % | 59±7 | 59±6 | 62±5 | 0.93 | 0.51 | 0.18 | 0.21 |
| LVM, g/m2 | 76±37 | 71±31 | 42±15 | 0.87 | **0.005** | **0.002** | **0.001** |
| GLS, % | -20.3±5.6 | -20.5±3.7 | -25.9±3.2 | 0.99 | **<0.001** | **<0.001** | **<0.001** |
| GCS, % | -30.3±8.7 | -29.8±5.8 | -34.7±5.8 | 0.98 | 0.14 | **0.026** | **0.029** |
| T1 Native, ms | 1040±55 | 985±43 | 972±29 | **0.007** | **<0.001** | 0.61 | **0.001** |
| ECV, % | 28.0±3.0 | 26.2±1.7 |  | **0.027** |  |  |  |
| T2, % | 53.6±1.6 | 50.6±2.5 | 50.7±2.0 | **0.002** | **0.001** | 0.99 | **0.006** |
| LGE positive, n(%) | 6 (50%) | 8 (25%) |  | 0.11 |  |  |  |

Abbreviations: E early diastolic mitral flow velocity, e' early diastolic tissue Doppler para-mitral segment velocity, A late ("atrial") diastolic mitral flow velocity, IVSd end-diastolic inter-ventricular septum thickness, LVIDd end-diastolic internal diameter, LVPWd end-diastolic posterior wall thickness, EDV end-diastolic volume, ESV end-systolic volume, SV stroke volume, CI cardiac index, EF ejection fraction, LVM left ventricular mass, GLS global longitudinal strain, GCS global circumferential strain, ECV extracellular volume, LGE late gadolinium enhancement.

**Figure 3.**
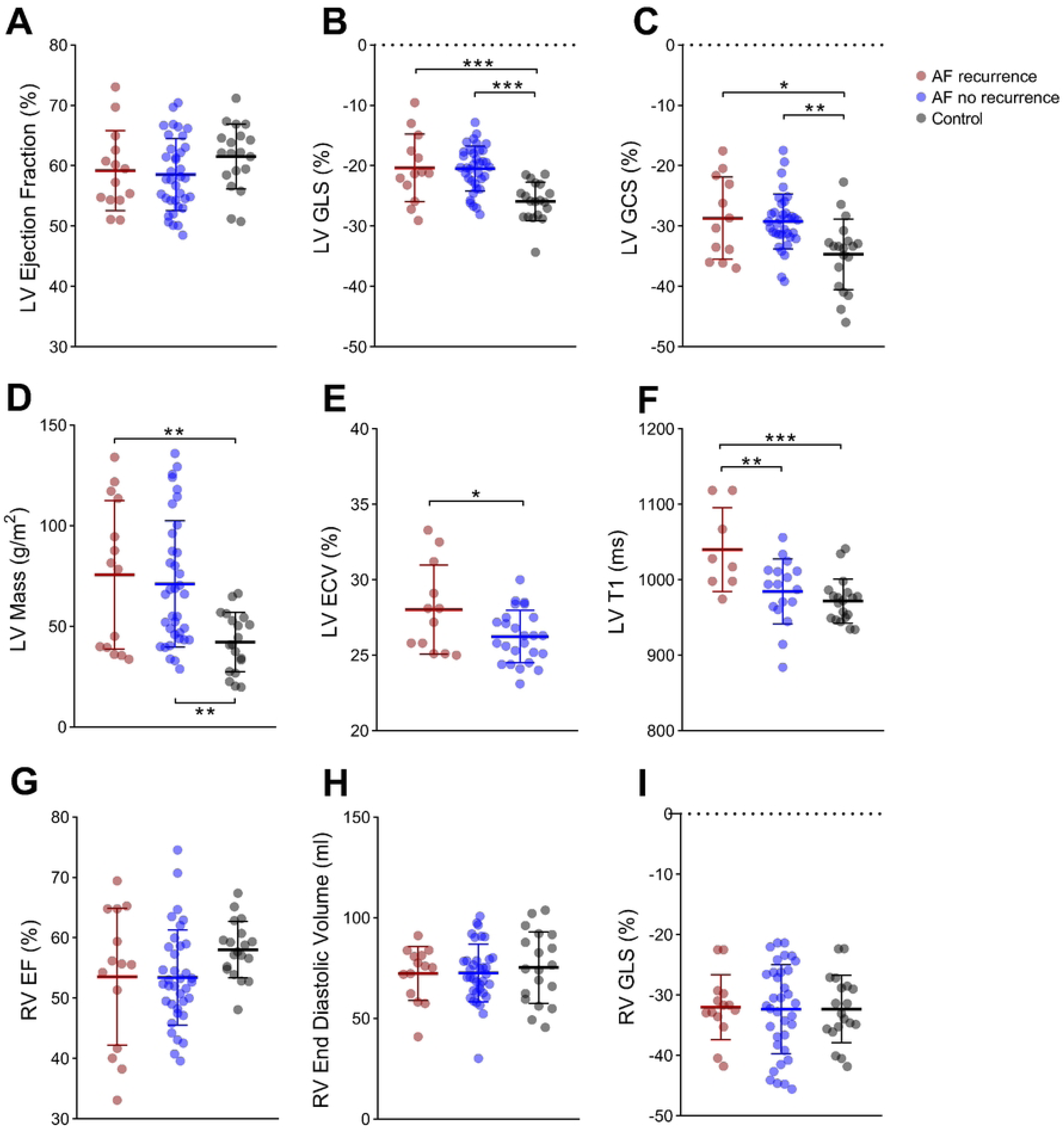
Cardiac remodelling post-RFCA: Left ventricle (Panels **A**, **B**, **C**, **D**, **E**, **F**): and the right ventricle (Panels **G**, **H**, **I**) across three groups: AF patients with recurrence post-RFCA (red dots), AF patients without recurrence post-RFCA (blue dots), and controls (black dots). LV ECV was not measured in Control group. While LV EF was not different between groups, LV Global Longitudinal Strain (**B**) and Global Circumferential Strain (**C**) were impaired in both sub-groups of AF patients and LV Mass (D), ECV (**E**) and T1 native (**F**) were higher in AF patients with recurrence. In the right ventricle, RV EF (**G**) was not different but Stroke Volume (**H**) was enlarged in both AF patient subgroups and RV Strain (**I**) was also not different overall. In all panels, parameters are presented as mean ± standard error, stratified by group. Significant differences between groups (p < 0.05) are indicated as follows: * p<0.05, ** p<0.01, *** p<0.001.

On echocardiogram, of note, there was no difference between groups in e’, E/e’, but A velocity was decreased in AF patients compared to controls and decreased also in AF-r compared with AF-nr (36.5±17.3cm/s vs 53.9±15.1cm/s vs 71.4±18.1cm/s, P_ANOVA_ < 0.001), in keeping with reduced atrial active contraction observed with CMR parameters.

### RV and RA remodeling

We further investigated the RV volumetry, function and strain and RA phasic volumes, function and strain (**Table 4**). There was no difference in RV volumes, EF or GLS between groups (**Figure 3 G, H, I**), but significantly more negative RV free-wall strain in AF-nr than in Control and a trend of more negative RV free-wall strain in AF-nr than in AF-r patients (−33.1±8.8% vs −39.1±8.3% vs −33.5±6.7%, P_ANOVA_= 0.016). There were no differences in RA phasic volumes, emptying fractions or strain between AF-r and AF-nr but there was a trend of larger volumes and more impaired emptying and strain in AF groups than Control. These reached significance for RA active emptying fraction (22±18% vs 28±17% vs 39±7%, P_ANOVA_= 0.007).

**Table 4.** Right Ventricular and Right Atrial Remodeling in 3 subgroups: AF patients with recurrence. AF patients with no recurrence and Control. Cardiac Magnetic Resonance Parameters.

|  | AF-recurrence<br>N=14 | AF-no recurrence<br>N=36 | Control<br>N=20 | P<br>AF rec vs non-rec | P<br>AF rec vs Control | P<br>AF non-rec vs Control | P<br>ANOVA |
| --- | --- | --- | --- | --- | --- | --- | --- |
| **<u>Right Ventricle</u>** |  |  |  |  |  |  |  |
| **CMR** |  |  |  |  |  |  |  |
| EDV | 72±13 | 73±14 | 75±18 | 0.99 | 0.85 | 0.81 | 0.80 |
| ESV | 32±9 | 30±8 | 32±9 | 0.81 | 0.99 | 0.77 | 0.72 |
| SV | 41±10 | 42±9 | 43±10 | 0.80 | 0.67 | 0.93 | 0.69 |
| EF | 54±11 | 53±8 | 58±5 | 0.99 | 0.26 | 0.11 | 0.12 |
| RV GLS | -32.0±5.4 | -32.4±7.4 | -32.3±5.6 | 0.98 | 0.99 | 0.99 | 0.98 |
| RV FW LS | -33.1±8.8 | -39.1±8.3 | -33.5±6.7 | 0.05 | 0.99 | **0.044** | **0.016** |
| **<u>Right Atrium</u>** |  |  |  |  |  |  |  |
| RA Volume Max | 55±21 | 57±18 | 50±16 | 0.94 | 0.67 | 0.34 | 0.37 |
| RA Volume Min | 36±19 | 33±15 | 25±8 | 0.77 | 0.07 | 0.11 | 0.05 |
| Reservoir RA EF | 36±17 | 43±14 | 50±8 | 0.23 | **0.010** | 0.15 | **0.013** |
| Reservoir RA Strain | 31.8±18.0 | 31.5±12.3 | 39.8±8.0 | 0.99 | 0.37 | 0.10 | 0.09 |
| Passive RA EF | 17.3±10.2 | 21.2±6.7 | 20.6±7.7 | 0.26 | 0.23 | 0.96 | 0.21 |
| Passive RA Strain | -17.2±12.5 | -15.3±5.3 | -19.2±4.3 | 0.69 | 0.71 | 0.15 | 0.17 |
| Active RA EF | 22±18 | 28±17 | 39±7 | 0.45 | **0.008** | **0.042** | **0.007** |
| Active RA Strain | -9.7±6.9 | -12.6±7.6 | -15.1±4.5 | 0.35 | 0.07 | 0.42 | 0.08 |

Abbreviations: EDV end-diastolic volume, ESV end-systolic volume, SV stroke volume, CI cardiac index, EF ejection fraction, GLS global longitudinal strain, FW LS free wall longitudinal strain, RA right atrial.

### AF recurrence and predictors

Next, we analysed which of the parameters characterising atrial and ventricular remodelling, alone or in combination, were predictive for AF recurrence. By testing the collinearity relation, LA Active Strain (AUC=0.774, P<0.001) was the most predictive parameter characterising LA remodelling. As T1 native, ECV and T2 were all higher in the AF-r than in AF-nr and showed only modest collinearity we included these three variables in a logistic regression model and performed ROC analysis to test the combined prediction ability of their combination (AUC= 0.871, P<0.001). When LA Active Strain was added to the model the AUC increased to 0.881, P<0.001. (**Figure 4 A**). Youden’s index (J) was estimated to determine the most predictive cut-off values, these were for LA Active Strain −8 %, for T1 native 1017.3 ms, for ECV 27% and for T2 52.4 ms. According to these cut-off values we further divided the AF patient group into two groups: Good LA Strain/Low LV ECV and Short T1 and T2 and, respectively Impaired LA Strain/ High LV ECV and Long T1 and T2 group. We compared these two groups regarding AF recurrence rate over 1-year follow-up. A Kaplan-Meier curve and Risk Table are presented in **Figure 4 B**. Patients in the Impaired LA Strain/ High LV ECV and Long T1,T2 group (N=16) are significantly more likely to have AF recurrence (HR: 17.70, 95% CI=3.89 - 80.61, log-rank P value <0.001) compared with patients in Good LA Strain/Low LV ECV and Short T1,T2 (N=34).

**Figure 4.**
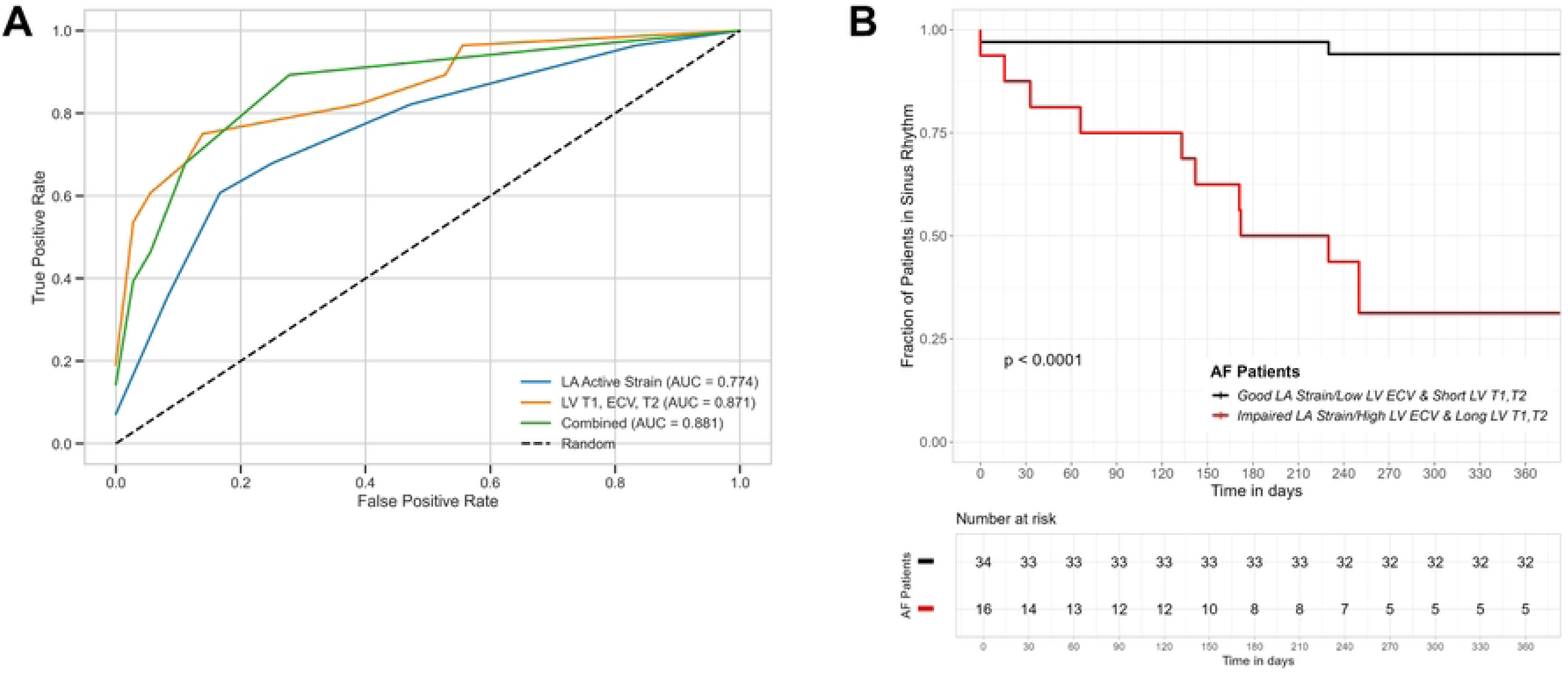
A. Receiver Operating Characteristic curves for LA Active Strain, T1 Native, LV Extracellular Volume (ECV), T2, and their combination. The ROC curve for LA Active Strain (AUC = 0.812) is shown in blue, while the ROC curve for the combined T1 Native, LV ECV, and T2 model (AUC = 0.854) is depicted in orange. The final combined model, integrating LA Active Strain with the synthetic variable, demonstrates superior predictive performance (AUC = 0.881), as shown in green. The dashed line represents the line of random discrimination (AUC = 0.5). Sensitivity and specificity were calculated at various thresholds to assess the discriminative ability of each model. **B.** Kaplan-Meier survival curves comparing groups defined by the combined variable from the ROC analysis. Patients were stratified into two groups: Group 1 (black line) – Low-risk group based on the combined variable, and Group 2 (red line) – High-risk group, based on the optimal threshold of the combined variable. The survival outcomes differ significantly between the two groups, as indicated by the log-rank test (p<0.001). The risk table below displays the number of patients at risk at each time point for both groups.

## Discussion

Given increasingly favourable data about mid and long-term prognosis upon CA ^26^, strategies to augment its long-term success in maintaining sinus rhythm remain a legitimate objective for electrophysiologists. However, current therapies still focus exclusively on isolating the ectopic electric activity from pulmonary veins and, in some cases non-pulmonary vein structures, yet ignore the complexities of cardiac atrial and ventricular remodelling concomitant to AF development. Using state-of-the-art CMR, our study findings add to the current knowledge on cardiac remodelling associated with AF after CA and its potential role for AF recurrence in patients without another significant cardiomyopathy and normal EF. Even if the EF remains within the normal range, both longitudinal and circumferential LV systolic strain were impaired in AF subjects. Parametric mapping further showed prolongated T1 native and T2 times in AF-r but not in AF-nr, and moreover ECV was increased in AF-r and AF-nr. Taken together, these findings indicate an important role of myocardial diffuse fibrosis and chronic inflammation in AF recurrence upon CA.

Frequently, a certain level of diffuse atrial fibrosis resulting from the activation of neurohumoral and other maladaptive pathways such as renin-angiotensin-aldosterone precedes the onset of AF. ^27^ Left untreated, AF itself further augments atrial structural remodelling leading to increase diffuse fibrosis, fatty infiltration, apoptosis and focal scarring and by that AF begets AF. ^28,29^ Thirdly, atrial fibrosis in AF patients with CA results from both the initial- and potential subsequent ablation lesions per se, and respectively from the hemodynamic consequences of AF. ^30^ Atrial cardiomyopathy and iatrogenic CA-related fibrosis ^31^ might determine a decrease in all three phasic atrial functions and mechanic deformation amplitude (strain). Indeed, it was previously shown quantitatively that the total amount of fibrosis is correlated tightly with atrial mechanic function, particularly with active emptying fractions and strain. ^32,33^ We showed here that an impaired AF active strain is associated with a higher risk of AF recurrence suggesting that this may indicate a more advanced atrial myopathy.

Post-procedural prevalence of thromboembolic events in AF patients undergoing CA is an important clinical outcome yet to be established. Mechanistically, the impact of AF CA on thromboembolic risk and on stroke prevalence remains elusive. Previous studies showed that reduced atrial strain in any of the 3 atrial phases of the cardiac cycle, with the tightest correlation observed for active LA strain, is associated with an increase in ischaemic stroke incidence in general population, effect maintained even in patients without atrial dilatation. ^34^ It can be speculated that impaired regional contraction, particularly in vulnerable areas, often subject to extensive ablation such as the pulmonary vein antrum or less dynamic regions close to the LA appendage have a higher prognostic role. This notion further supports the currently suggested initiated and/or (transiently) maintained oral anticoagulation after CA.

We found that, despite maintaining a normal EF after ablation, patients with AF undergoing CA had impaired myocardial systolic LV GLS and GCS with no difference between AF-r and AF-nr. This data points towards a subtle ventricular mechanic impairment in our AF cohort. A higher prevalence of coronary artery disease ^35^ and a pre-existent metabolic deficit at the myocardial level ^36^ have been found among patients with AF, both of which would cause contractile impairment. A reduced atrial contribution to diastole, increasingly important in older individuals, may also explain worsening myocardial perfusion, leading to decreased ventricular contractility. ^37^

The impact of AF ablation on RV function and the pulmonary vasculature in patients with normal EF and no other known cardiac pathology is currently incompletely explored. RV function mirrors the sum of increased LV diastolic filling pressures, atrial reservoir stiffness and reduced pulmonary vasculature compliance and increased resistance. ^38^ While successful ablation often improves atrioventricular synchrony, reduces atrial strain, and may lead to reverse RV remodelling, persistent RV dilation can occur in some cases. ^39^ We tested this hypothesis by quantifying the RV volumes, RV global and free-wall strain. There were no significant differences between groups and, paradoxically, more negative RV free-wall strain in AF-nr. This incidental finding will need to be confirmed in a larger number of patients but can indicate the importance of contractile RV reserve in compensating for a reduced LA reservoir function.

We found that all T1 native, ECV and T2 are elevated in patients with AF-r and are not different from control in patients with AF-nr and successful CA. These data emphasize the importance and magnitude of underlying myocardial pathology in AF pathophysiology and severity. An increased level of ventricular diffuse fibrosis and inflammation were observed in patients with AF-r but not in those where SR was successfully maintained at 1-year follow-up. Moreover, we showed here that combining LV T1 native, ECV and T2 parameters adds to the prognostic role of LA Active Strain with regard to AF recurrences, offering a powerful tool to risk stratify AF recurrencies post CA. Due to the current absence of specific atrial tissue mapping options, the question remains open for further investigations, if similar levels of structural changes are present at the atrial level and, in fact, if those are responsible for more electrical instability and AF re-emergence.

## Limitations

This is a cross-sectional study and therefore we could not establish the dynamic of atrial and ventricular remodelling with a long-term follow-up after CA procedure. Particularly, a more definite improvement of atrial function and a regression of RV congestion are important factors, that remain to be established by a prolonged survey of these patients. This favourable response can also have prognostic implications for long-term maintenance of SR. Finally, we used the biplane area-length method to evaluate atrial phasic volumes and emptying fraction. However, even more precise methods such as a multi-disc stack evaluation may yield better accuracy in assessment of atrial volumes, function and strain.

## Data Availability

All relevant data are within the manuscript and its Supporting Information files.

## List of abbreviations

AF: Atrial fibrillation
CA: Catheter ablation
CMR: Cardiac magnetic resonance
LGE: Late gadolinium enhancement
LA: Left atrium
LV: Left ventricle
RV: Right ventricle
RFCA: Radiofrequency catheter ablation
SR: Sinus rhythm
ACT: Activated clotting time
SCMR: Society for Cardiovascular Magnetic Resonance
SSFP: Steady-state free precession
MOLLI: Modified Look-Locker Inversion recovery
GraSE: Gradient and spin echo
EPI: Echo planar imaging
SENSE: Sensitivity encoding
FT: Feature tracking
OP: Outpatient
EA: Ethics approval
TE: Echo time
TR: Repetition time
FA: Flip angle

## Clinical Perspective

Our data contribute important insights to the paradigm that AF is a condition involving complex underlying changes in cardiac structures, which are both causal and consequential to the presence and progression of arrhythmic events. More efficient clinical care following AF ablation could potentially be achieved by considering the degree of pre-procedural cardiac remodelling and monitoring its potential reversal post-procedure.

